# A feasibility trial of a digital mindfulness-based intervention to improve asthma-related quality of life for primary care patients with asthma

**DOI:** 10.1101/2021.04.27.21256158

**Authors:** Ben Ainsworth, Sabina Stanescu, Beth Stuart, Daniel Russell, Megan Liddiard, Ratko Djukanovic, Mike Thomas

## Abstract

**Objective:** Asthma outcomes remain suboptimal, despite effective pharmacotherapy. Psychological dysfunction (such as anxiety) is common, and associated with poorer outcomes. We evaluated a digital mindfulness programme as an intervention to improve asthma-related quality of life for primary care patients, in a prospectively registered randomized-controlled feasibility study.

**Methods:** We offered ‘Headspace’, a widely-used digital mindfulness intervention, to adults with asthma through 16 UK GP practices. Participants were randomised on a 2:1 basis to the mindfulness intervention, or waitlist control. Participants completed questionnaires (including asthma symptom control, asthma-related quality of life, anxiety, depression) at baseline, 6-week and 3-month follow-up.

**Results:** 114 participants completed primary outcomes at 3-month follow-up (intervention 73 (71.6%), control 41 (70.7%)). Compared to baseline, the intervention group but not the control group reported significantly improved asthma-related quality of life, with a non-significant between-group difference favouring the intervention group (Mean difference = 0.16, 95%CI: - 0.11 – 0.44). Intervention use varied but was generally high.

**Conclusions:** Digital mindfulness interventions are feasible and acceptable adjunct treatments for mild and moderate asthma to target quality of life. Further research should adapt ‘generic’ mindfulness-based stress-reduction to maximize effectiveness for asthma, and validate our findings in a fully-powered randomized controlled trial.

**Trial registration:** Prospectively registered: ISRCTN52212323

---

Asthma is a multifaceted chronic disease, with recent estimates that it affects 339 million people of all ages worldwide and 6.5% of the UK population (Bloom et al., 2019). Although evidence suggests that modern pharmacotherapy can achieve good asthma control in clinical trials (Bateman et al., 2004), in reality the heterogenous clinical and behavioural phenotypes mean that asthma outcomes remain suboptimal, and many patients continue to experience persistent symptoms and impaired quality of life (Demoly et al., 2010).

The causes of these suboptimal therapeutic outcomes are complex and wide-ranging, including poor self-management (ie corticosteroid inhaler adherence and technique; see Thomas, 2015 for a review), but increasingly the role of psychological comorbidity including anxiety, depression and panic has become apparent (Gada et al., 2014; Goldney et al., 2003; Hasler et al., 2005; Shaw et al., 2015). Anxiety and depression-related psychological dysfunction are up to six times more common in people with an asthma diagnosis (Goodwin et al., 2003; Lavoie et al., 2006), and even more likely with difficult-to-control asthma (Lavoie et al., 2006). With frequent experiences of unpredictable and potentially life-threatening breathlessness, psychological dysfunction is also associated with avoidant coping strategies (leading to lower quality of life; (Adams et al., 2004; Cluley and Cochrane, 2001) and increased healthcare utilisation (Richardson et al., 2008). Recent reviews have highlighted the need for appropriate treatment that considers these psychological aspects that will improve patient well-being and asthma control (Baiardini et al., 2015).

Current research in asthma suggests that behavioural and psychological interventions that aim to improve health outcomes can potentially be effective, although the quality and volume of research performed, the variety of interventions investigated(including relaxation, biofeedback, mindfulness and self-management) and the variety of health-related outcome measures reported, means that evidence for specific treatments generally remains inconclusive (Yorke et al., 2015). However, a recent large-scale randomised controlled trial (RCT) found that self-guided breathing exercises for asthma were effective and cost-effective to improve quality of life (Bruton et al., 2017), and this intervention is now advocated in evidence-based asthma guidelines (James and Lyttle, 2016; Reddel et al., 2015).

Mindfulness meditation-based interventions (MBIs) can potentially offer benefit for people with asthma, although as with other non-pharmacological treatments evidence is inconclusive (Paudyal et al., 2018). Mindfulness-based stress reduction (MBSR) and mindfulness-based cognitive therapy (MBCT) are common treatments for anxiety and depression (Strauss et al., 2014) and have demonstrated benefit across a range of chronic conditions (eg. fibromyalgia, cancer, arthritis and cardiovascular disease; see (Bohlmeijer et al., 2010). Cross-sectional studies have found that higher trait mindfulness is associated with reduced asthma symptoms (Kraemer and McLeish, 2019; Shi et al., 2018), and participants randomised to an 8-week MBSR course (vs. psychoeducation) showed improved quality of life and perceived stress (Pbert et al., 2012).

A common barrier to the implementation of MBIs in chronic disease is the burden of attending the weekly group sessions – for example, a standard MBSR course might consist of 8 two-hour sessions once per week, with additional self-practice (Ainsworth et al., 2020; Simpson et al., 2018). MBIs are complex behavioural interventions and it is therefore possible that they may require innovative delivery models to maximise access and effectiveness across different patient groups and to achieve cost-effectiveness (Demarzo et al., 2015). Digital mindfulness interventions could potentially offer alternatives to traditional programmes, allowing accessibility to content that has been created and validated by experts, across a heterogenous population at low-cost. Digital self-management support interventions have been successfully trialled in asthma, with patient acceptability (Deborah Morrison et al., 2014; Ainsworth et al, 2019a). Web-based MBIs have shown some benefit in alleviating symptom burden across non-respiratory chronic conditions (Toivonen et al., 2017), but to date a digital mindfulness intervention has not been evaluated for adults with asthma.

This study aimed to explore the feasibility of using ‘Headspace’, a market-leading digital mindfulness intervention that is commercially available (Mani et al., 2015) for improving patient reported outcomes for people with mild and moderate asthma treated in primary care, and to estimate effect size for a subsequent fully powered trial.

## Methods

### Objectives

Specific study objectives were to:

1. Explore recruitment procedures including rates of invitation response, study recruitment, randomisation and retention in a feaslibility pilot randomised controlled trial of a digital mindfulness intervention for people with asthma.
2. Describe and evaluate changes in of baseline and 3-month follow-up self-report measures of quality of life, asthma control, anxiety and depression.
3. Examine intervention usage and engagement to inform a future modified intervention.

### Design

The study was a prospective randomized-controlled feasibility trial comparing free access to the digital mindfulness programe with usual care, using simple randomisation (weighted: 2 intervention vs. 1 control, in order to examine intervention usage and engagement in depth).

Ethical approval was given by the South Central Hampshire Research Ethics Committee: 17/SC/0088. No formal power calculation was conducted as this was a feasibility study.

### Participants

Participants meeting the inclusion criteria were identified in searches of GP electronic clinical records and invited from practices in Hampshire, UK. Target sample size was 120 (80 intervention, 40 control), considered sufficient to explore study feasibility and intervention acceptability.

Inclusion criteria: Over 18 years old, clinical asthma diagnosis and current treatment in primary care (confirmed by one or more asthma medication prescription in previous year).

Exclusion criteria: previous diagnosis of major or unstable comorbid psychological disorders, other than anxiety or depression, currently participating in another asthma interventional study, acute exacerbation of asthma requiring a course of oral steroids within previous 28 days, asthma treated in secondary care.

Recruitment was conducted from July 2017 to April 2018.

### Outcome measures

The study and measures were prospectively registered on the ISRCTN Registry (reference 52212323). The main outcome of the study was the feasibility of trial procedures (recruitment and randomisation rates, intervention engagement, completion and acceptability of outcome measures).

#### Self-report and clinical measures

Alongside feasibility and recruitment measures, the main study outcome of interest was asthma-related quality of life, measured using the mini-Asthma Quality of Life Questionnaire [AQLQ:(Juniper et al., 1992)], a validated 15-item questionnaire in which participants assess their asthma-related wellbeing over the last two weeks. The overall score is the mean of all items (7 = not impaired at all, 1 = severely impaired), with 4 subscales (symptoms, activities, emotion, environment) with a minimum clinically important difference (MCID) for individual patients of 0.5. A higher score equates to better quality of life. Baseline scores demonstrated good internal reliablity (α = 0.92).

Asthma control was measured with the 6-item Asthma Control Questionnaire [ACQ: (Juniper et al., 1999)], with a lower score equating to improved asthma control (α = 0.88). Anxiety and depression were measured with the Hospital Anxiety and Depression Scale [HADS: (Zigmond and Snaith, 1983); anxiety α = 0.87, depression α = 0.84]. Mindfulness was measured with the Philadelphia Mindfulness Scale [PHLMS: (Cardaciotto et al., 2008); awareness α = 0.83; acceptance α = 0.88]. Medication adherence was measured with the Medical Adherence Report Scale - Asthma [MARS-A: (Mora et al., 2011); α = 0.83].

Participants responding to the study invitation completed online consent and confirmed eligibility on the computerized Lifeguide platform (Yardley et al., 2009) before completing baseline demographic and self-report measures of anxiety, depression, mindfulness and medication adherence. The primary outcome (AQLQ) was completed via post (as it was not available for on-line completion), as was the ACQ.

Participants were randomised by the Lifeguide software and those allocated to the intervention were sent intervention access instructions via email and post. Six week and three month follow-up measurements were conducted via post (asthma quality of life, asthma control) and online (anxiety, depression, mindfulness and medication adherence).

Additional exploratory outcome measures of participant enablement, acceptance and action and illness perceptions were also recorded, alongside qualitative interviews, for a detailed process analysis that will be reported in a separate paper.

### Intervention

#### Mindfulness Intervention

The commercially-available Headspace app (‘Meditation and Sleep Made Simple—Headspace’, available from http://www.headspace.com/) In an empirical examination of the quality of 23 commercially-available meditation apps, Headspace was rated the highest based on various criteria including engagement, functionality and information quality (Mani et al., 2015). It is available on iOS and Android smartphones, and desktop computers. Appendix 1 describes Headspace according to the template for intervention description and replication (TIDieR; Hoffmann et al., 2014).

While Headspace does contain brief online written information (e.g. ‘the science of meditation’) the primary content is 3, 5, 10 and 20 minute long audio-guided meditations. The Headspace app contains more than 200 different courses covering a wide range of topics, from stress and mental health to physical health, job performance, and emotional well-being. Individuals new to mindfulness and meditation can learn the fundamentals by engaging with the three “basics” courses, each consisting of ten guided meditations. Upon randomization to the intervention group (or after completing final follow-up if randomized to the waitlist control group), participants were provided with unique redemption codes providing them with 6 months of free access to the complete Headspace content library. Participants were sent standard instructions on how to activate their Headspace accounts using their free access code via email and post.

### Recruitment, retention and adherence

Participant recruitment was conducted with the NIHR Clinical Research Network (CRN) Wessex, who contacted local GP practices to confirm interest. Staff across 17 GP practices (mean list size 9584) across Hampshire searched patient records, identifying 6243 patients meeting eligibility criteria (see Figure 1). 4401 patient records were screened by clinicians before being contacted with a letter from their GP and a copy of the participant information sheet. Participants were invited to contact the study team to express interest, or could sign up using the study website.

**Figure 1:**
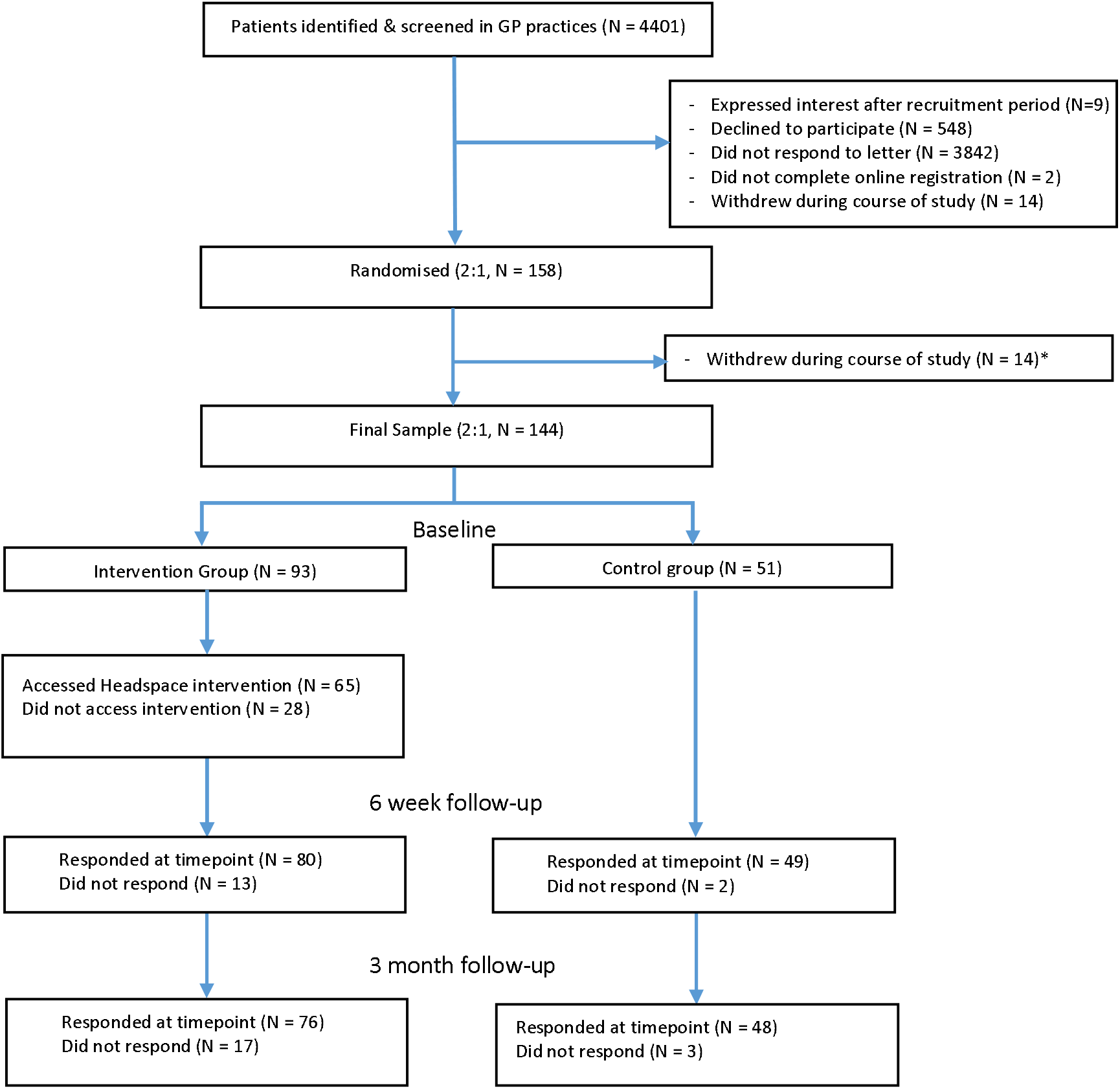
Consort Diagram of recruitment and retention during study. **Note:** Participants completed both online and postal measures and are included here if they completed either. A full breakdown of the measures completed by participants at each timepoint (and therefore how many patients were available for subsequent analysis) is available in Table 2. **Reasons for withdrawal:** were loss of interest (N = 4), lack of time (N = 2), personal issues/illness not related to study (N = 3), wanted to use intervention despite being in control group (N = 1), difficulty using

A total of 158 participants provided informed consent and were randomised after completing baseline questionnaires. Intervention group participants were granted access to the intervention immediately while control group participants were told they would receive access after they had completed further questionnaires in six weeks and three months. 14 participants (5 control, 9 intervention) withdrew during the course of the study leaving a final sample of 144 (control 51 / intervention 93). Reasons for withdrawal are reported in the Consort diagram (Figure 1). No harms were reported during the study.

548 participants (37% male, age M = 62) returned optional opt-out postal slips with free-text reasons for opting out. Common reasons for opting out included considering asthma as not severe enough (N = 82) or well-controlled (N = 43), no asthma symptoms (N = 20), not having asthma (N = 23), not having access to the internet (N = 82), not interested in meditation (N = 9), already experienced with meditation (N = 12) or too busy to take part (N = 9).

A full CONSORT flow diagram of the study is presented in Figure 1.

### Analysis

Study outcome data were examined using using SPSS v24 and the results are presented descriptively. Independent group comparisons compared baseline differences. Within-group and between-group changes from baseline in the key outcome measures were assessed, including estimations of proportions reaching the minimum clinically important differences in the AQLQ (MCID: 0.5). The feasibility study was not powered to detect significant differences from pre-test to post-test or between groups, but exploratory comparisons using linear regression, controlling for baseline values of each outcome measure, examined changes in self-report measures between groups to inform future power calculations. Missing data was not imputed and the results presented here represent complete cases only.

## Results

### Participants

Participants had a large range of education, internet experience, meditation experience and time since asthma diagnosis, and were predominantly white. Baseline demographic and outcome measures scores, collected online during study registration, are reported in more detail in Table 1.

**Table 1.** Baseline participant demographic information and outcome scores for intervention and control groups.

| *Measure* | **Intervention (N = 93)** | **Control (N = 51)** | **Mean difference (95% CI)** |
| --- | --- | --- | --- |
| Age: M (SD) | 49.8 (14.7) | 53.5 (14.4) | 3.7 (-1.4, 8.8) |
| Ethnicity (%) | White (97%), Indian (3%), | White (93%), Chinese/South East Asian (2%), Indian (2%), Other (2%) | - |
| Education (%) | School (22%), Degree/Diploma (57%), Postgraduate (20%), Other (1%) | No formal (5%), School (52%), Degree/Diploma (26%), Postgraduate (14%), other (2%) | - |
| Weekly internet use: M hours (SD) | 17.6 (14.4) | 16.4 (14.3) | -1.3 (-6.9, 4.3) |
| Meditation experience (%) | Not heard of it/don't know about it (14%), never tried it (30%), tried other types of meditation (23%), tried mindfulness (25%), regularly practice mindfulness (7%). | Not heard of it/don't know about it (24%), never tried it (37%), tried other types of meditation (17%), tried mindfulness (16%), regularly practice mindfulness (5%). | - |
| *Years since diagnosis* | 28.2 (15.3) | 21.1 (16.2) | -6.8 (-13.0, -0.6)* |
| *Asthma-related Quality of Life (AQLQ)* | 5.32 (1.1) | 5.64 (1.0) | 0.32 (-0.05, 0.69) |
| *Symptoms subdomain* | 5.09 (1.2) | 5.52 (1.2) | 0.43 (0.01, 0.87)* |
| *Environment subdomain* | 4.92 (1.4) | 5.05 (1.4) | 0.13 (-0.35, 0.62) |
| *Emotions subdomain* | 5.26 (1.4) | 5.76 (1.3) | 0.51 (0.02, 0.99)* |
| *Activities subdomain* | 5.85 (1.1) | 6.01 (0.9) | 0.17 (-0.20, 0.54) |
| *Asthma Control (ACQ)* | 1.18 (0.9) | 1.06 (0.8) | -0.12 (-0.43, 0.18) |
| *Anxiety (HADS-A)* | 8.24 (4.3) | 6.76 (4.1) | -1.47 (-2.91, -0.01)* |
| *Depression (HADS-D)* | 4.72 (3.9) | 3.65 (3.1) | -1.07 (-2.33, 0.19) |
| *Mindful Awareness (PHLMS-Aw)* | 31.3 (7.7) | 29.6 (8.1) | -1.74 (-4.45, 0.97) |
| *Mindful Acceptance (PHLMS-Ac)* | 35.1 (6.8) | 33.9 (5.7) | -1.26 (-3.46, 0.95) |
| *Medication Adherence (MARS-A)* | 37.9 (8.5) | 38.1 (8.4) | 0.25 (-2.68, 3.18) |

**Note:** ACQ (lower scores equate to better control); AQLQ (higher scores equate to greater impairment); HADS (higher scores equate to more anxiety). PHLMS (two subscales of awareness and attention, in which higher scores equate to more mindfulness); MARS-A (higher scores equate to better adherence). (\*) denotes group differences in which the 95% CI does not include 0.

Baseline group comparisons showed some imbalances between the randomisation groups: people randomised to the intervention group were diagnosed longer ago, and had greater impairment on AQLQ symptom and emotion subdomain, higher anxiety scores, greater overall AQLQ impairment and lower asthma control scores. Our primary analysis of follow-up comparisons therefore controlled for baseline differences in each measure by including them as covariates within the regression model.

### Intervention engagement

65 participants in the intervention group (70%) accessed the app through the access code provided on one or more occasions, with a total of 2478 recorded individual sessions. Usage after the 3 month trial period had ended was not included in analysis.

Participants in the intervention group accessed the intervention between 0 and 192 times each (Median 9.0, IQR 0-38.5). 28 participants did not access the intervention at all. Average session length was 7.2 minutes (SD 3.0) with 682 different practice recordings used across 232 session types. The mean number of days between the first and last use (during the trial) was 51 days.

Participants most frequently accessed the initial introductory session (‘Basics’, Median 6.6 times accessed, Range 13, accessed by 100% of users) as well as ‘Managing Anxiety’ (M 4.0, R 31, 31% user access), Basics 2 (M 3.0, R 13, 55%) and Basics 3(M 2.6, R 11, 33%). Only the most popular sessions (introductory and anxiety management) were used consistently across participants; most other sessions (eg. stress, self-esteem, sleep) were used by fewer than 5 individuals.

Exploratory analysis allocated participants into users who were non-engagers (0 log ins, N = 28), low engagers (1-5 sessions, N = 14), moderate engagers (6-15 sessions, N = 17), high engagers (16-50 sessions, N = 16) and very high engagers (50+ sessions, N = 18). Baseline group comparisons found no differences in baseline scores of asthma control, asthma-related quality of life, depression, mindfulness or medication adherence (ps > .10). Groups differed in anxiety (F_(4,95)_ = 4.69, p = .002) with low anxiety scores in non-engagers (M 6.89, 95% CI 5.26-8.53) and high engagers (M 6.06, 95% CI 3.93-8.20) compared to low engagers (M 11.21, 95%CI 9.03-13.40), moderate engagers (M 8.24, 95% CI 6.09-10.38) and very high engagers (M 9.94, 95% CI 7.35-9.13).

### Outcome measure response rate

Total responses to outcome measures are reported in a consort diagram in Figure 1. Most participants completed both online and postal questionnaires, with similar completion in both groups (see Table 2). Completion rates were slightly higher in the control group. Further analyses at each time point included all participants who completed data for that timepoint (for example, participants who completed measures at 3-months but not 6-weeks were still included in the 3-month followup analysis).

**Table 2.** Postal and Online questionnaire response rates.

| *Measures completed* | **Intervention N (%)**<br>**(Randomised N = 93)** |  |  | **Control N (%)**<br>**(Randomised N = 51)** |  |  |
| --- | --- | --- | --- | --- | --- | --- |
|  | **Baseline** | **6-week** | **3-month** | **Baseline** | **6-week** | **3-month** |
| *Both postal and online (all measures)* | 84 (90) | 50 (54) | 62 (67) | 48 (94) | 38 (75) | 38 (75) |
| *Postal only (AQLQ, ACQ)* | 0 (0) | 15 (16) | 11 (12) | 0 (0) | 8 (16) | 5 (10) |
| *Online Only (HADS, PHLMS, MARS-A)* | 9 (10) | 15 (16) | 3 (3) | 3 (6) | 3 (6) | 5 (10) |
| *None* | 0 (0) | 13 (14) | 17 (18) | 0 (0) | 2 (4) | 3 (6) |
| *Primary Endpoint Measure (AQLQ)* | 84 (90) | 65 (70) | 73 (79) | 48 (94) | 46 (91) | 43 (85) |

**Note:** Participants who actively withdrew from study were excluded from this analysis, those lost to follow-up were not.

### Primary Analysis: Asthma Quality of Life Questionnaire and Asthma Control Questionnaire

Follow-up scores are reported in Table 3, with scores for each measure at each time-point, and group comparisons of estimated marginal means (ie. between-group comparisons corrected for baseline differences, reported in Table 2) at 6-week and 3-month. Between group comparisons were followed by comparisons within each group (baseline vs. follow-up).

**Table 3.** Baseline, 6- week and 3-month follow-up questionnaire scores of randomised intervention (N=93) vs. control participants (N=51).

| Postal Measures | Intervention (M, SD) |  |  | Control (M, SD) |  |  | Intervention vs. Control Comparison (M, 95% CI) |  |
| --- | --- | --- | --- | --- | --- | --- | --- | --- |
|  | Baseline<br>(N = 84) | 6-week<br>(N = 64) | 3-month<br>(N = 73) | Baseline<br>(N = 48) | 6-week<br>(N = 46) | 3-month<br>(N = 43) | Baseline vs. 6-week<br>(N = 63 vs 45) | Baseline vs. 3-month<br>(N = 71 vs 42) |
| *Asthma-related Quality of Life (AQLQ)* | 5.32 (1.1) | 5.67 (1.0) | 5.77 (0.9) | 5.64 (1.0) | 5.72 (1.0) | 5.77 (0.9) | 0.20 (-0.06, 0.46) | 0.15 (-0.13, 0.42) |
| *Symptoms subdomain* | 5.09 (1.2) | 5.56 (1.1) | 5.60 (1.1) | 5.52 (1.2) | 5.60 (1.1) | 5.60 (1.1) | 0.25 (-0.08, 0.59) | 0.15 (-0.21, 0.51) |
| *Environment subdomain* | 4.92 (1.4) | 5.33 (1.1) | 5.58 (1.2) | 5.05 (1.3) | 5.30 (1.4) | 5.58 (1.3) | 0.10 (-0.24, 0.45) | 0.26 (-0.08, 0.60) |
| *Emotions subdomain* | 5.26 (1.4) | 5.57 (1.4) | 5.74 (1.2) | 5.76 (1.3) | 5.86 (1.3) | 5.74 (1.1) | 0.08 (-0.32, 0.49) | -0.11 (-0.45, 0.23) |
| *Activities subdomain* | 5.85 (1.1) | 6.15 (1.0) | 6.14 (1.0) | 6.02 (0.9) | 6.07 (1.1) | 6.14 (1.0) | 0.20 (-0.09, 0.48) | 0.20 (-0.11, 0.51) |
| *Asthma Control (ACQ)* | 1.18 (0.9) | 1.02 (0.9) | 1.00 (0.8) | 1.08 (0.8) | 1.14 (0.8) | 1.15 (0.9) | -0.26 (-0.49, -0.03)* | -0.17 (-0.44, 0.10) |
| Online Measures | Baseline<br>(N = 93) | 6-week<br>(N = 65) | 3-month<br>(N = 65) | Baseline<br>(N = 51) | 6-week<br>(N = 41) | 3-month<br>(N = 43) | Baseline vs. 6-week<br>(N = 65 vs. 41) | Baseline vs. 3-month<br>(N = 65 vs. 43) |
| *Anxiety (HADS-A)* | 8.24 (4.3) | 8.58 (3.3) | 7.46 (3.5) | 6.76 (4.1) | 7.78 (2.8) | 7.40 (3.0) | -0.18 (-1.00, 0.63) | -0.62 (-1.50, 0.27) |
| *Depression (HADS-D)* | 4.72 (3.9) | 3.86 (3.6) | 3.49 (3.3) | 3.65 (3.1) | 4.15 (3.8) | 4.30 (3.5) | -1.34 (-2.29, -0.39)* | -1.63 (-2.48, -0.77)* |
| *Mindful Awareness (PHLMS-Aw)* | 31.3 (7.7) | 29.1 (7.5) | 26.4 (8.4) | 29.6 (8.1) | 28.4 (6.5) | 27.1 (8.1) | 0.76 (-1.39, 2.91) | 1.74 (-0.55, 4.03) |
| *Mindful Acceptance (PHLMS-Ac)* | 35.1 (6.7) | 36.0 (6.1) | 36.8 (5.9) | 33.9 (5.7) | 33.5 (5.6) | 33.3 (7.3) | 1.31 (-0.45, 3.07) | 1.63 (-0.41, 3.67) |
| *Medication Adherence (MARS-A)* | 37.9 (8.5) | 39.5 (9.3) | 39.8 (9.7) | 38.1 (8.4) | 38.4 (8.9) | 37.7 (9.0) | 0.16 (-2.16, 2.48) | 1.39 (-0.91, 3.68) |

**Notes:** Between group differences are reported as estimated marginal mean difference scores (corrected for baseline values of each measure). Missing data was not imputed and data presented here represent a modified intention to treat analysis, with participants analysed as randomised but only if they completed the follow up measures. (\*) denotes differences in which the 95% confidence interval of group differences does not contain 0. As participants were able to complete postal or online measures, Ns for each analysis have been reported separately.

#### Asthma Quality of Life Questionnaire (AQLQ)

##### 6-week follow-up

Between group comparison (correcting for baseline differences) showed higher AQLQ scores in the the intervention group above the control group (Mean Difference [MD] 0.20, 95%CI -0.06-0.46) although this difference was not significant. Within-group analysis from baseline to 6-weeks showed significantly improved AQLQ score in the intervention group (MD 0.34, 95%CI 0.15, 0.52), but not in the control group (MD 0.03, 95%CI - 0.19, 0.24).

##### 3-month follow-up

Between group (correcting for baseline differences) showed improved AQLQ scores in the intervention group in comparison with the control group (MD 0.15, 95%CI -0.13, 0.42), although as at 6 weeks this different was not significant. As with the 6-week analysis, within-group mean AQLQ score changes from baseline in the intervention group significantly improved (MD 0.39, 95%CI 0.18, 0.59) but not in the control group AQLQ (MD 0.11, 95%CI -0.13, 0.36).

#### Asthma Quality of Life Subscales: Symptoms (AQLQ-S), Environment (AQLQ-En), Emotion (AQLQ-En), Activities (AQLQ-A)

##### 6-week follow-up

Between group comparisons (correcting for baseline differences) showed improvements in the intervention group in subdomain scores of symptoms (MD 0.25, 95%CI - 0.08, 0.59), environment (MD 0.10, 95%CI -0.24, 0.45), emotions (MD 0.08, 95%CI -0.32, 0.49). and activities (MD 0.20, 95%CI -0.09, 0.48) but these were not significant.

Within group comparisons from baseline to 6-weeks showed the intervention group improved significantly in all subdomains (Symptoms MD 0.48, 95%CI 0.24, 0.73; Environment MD 0.37, 95%CI 0.10, 0.64; Emotion MD 0.33, 95%CI 0.06, 0.59; Activities MD 0.26, 95%CI 0.07, 0.45) while the control group did not significantly improve in any subdomains (Symptoms MD 0.02, 95%CI -0.28, 0.32; Environment MD 0.21, 95%CI -0.06, 0.50; Emotion MD 0.04, 95%CI -0.32, 0.41; Activities MD 0.01, 95%CI -0.23, 0.26).

##### 3-month follow-up

Between group comparisons (correcting for baseline) showed non-significant improvements across subdomain scores of symptoms (MD 0.15, 95%CI -0.21, 0.51), environment (MD 0.26, 95%CI -0.08, 0.60) and activities (MD 0.20, 95%CI -0.11, 0.51) in the intervention group compared to the control group, but not in emotions (MD -0.11, 95%CI -0.45, 0.23).

Within group comparisons from baseline to 3-months showed the intervention group improved significantly in all subdomains (Symptoms MD 0.43, 95%CI 0.17, 0.69; Environment MD 0.62, 95%CI 0.39, 0.85; Emotion MD 0.41, 95%CI 0.14, 0.68; Activities MD 0.25, 95%CI 0.04, 0.46) while the control group did not significantly improve in any subdomains (Symptoms MD 0.07, 95%CI -0.26, 0.39; Environment MD 0.30, 95%CI -0.04, 0.63; Emotion MD 0.31, 95%CI -0.004, 0.62; Activities MD 0.02, 95%CI -0.23, 0.23).

#### Asthma Control Questionnaire (ACQ)

##### 6-week follow-up

Between group comparisons (correcting for baseline) showed the ACQ score improved in the intervention group above the control group (MD -0.26, 95%CI -0.49, -0.03; with a lower ACQ score indicating improved control). Within group comparisons showed that the intervention group had significantly improved mean ACQ vs. baseline (MD -0.17, 95%CI -0.33, - 0.01) while asthma control in the control group reduced although not significantly (MD -0.13, 95%CI -0.06, 0.31).

##### 3-month follow-up

Between-group comparisons (correcting for baseline) showed improvement in ACQ score in the intervention group above the control group (MD -0.17, 95%CI -0.44, 0.10) but this was not significant. Within-group comprison from baseline to 3-months improvement in the intervention group (MD -0.12, 95%CI -0.32, 0.07) and worsening in the control group (MD 0.10, 95%CI -0.12, 0.32) which were both non-significant.

### Secondary analysis: Anxiety, Depression, Mindfulness and Medication Adherence

Additional outcomes of anxiety (HADS-A), depression (HADS-D), mindfulness (PHLMS-Aw and PHLMS-Acc) and medication adherence (MARS-A) were examined using group comparisons at follow-up (correcting for baseline differences in each measure). Data from both 6-week and 3-month follow-up is reported in full in Table 3.

At 6-week followup, the intervention group had significantly lower depression scores at follow-up compared to the control group (MD -1.34, 95%CI -2.29, -0.39). The intervention group also had lower anxiety scores (MD -0.18, 95%CI -1.00, 0.63), higher mindful awareness (MD 0.76, 95%CI -1.39, 2.91) and higher mindful acceptance (MD 1.31, 95%CI -0.45, 3.07) than the control group, but these differences were not significant. There were no differences in medication adherence (MD 0.16, 95%CI -2.16, 2.48).

The same pattern was observed at 3-month follow-up. In comparison with the control group, the intervention group had significantly lower depression scores (MD -1.63, 95%CI -2.48, -0.77), and lower (but not significantly so) scores in anxiety (MD -0.62, 95%CI -1.50, 0.27), awareness (MD 1.74, 95%CI -0.55, 4.03), and acceptance (MD 1. .63, 95%CI -0.41, 3.67). The intervention group also demonstrated slightly better adherence scores (MD 1.39, 95%CU -0.91, 3.68) although this difference was also not significant.

Within-groups analysis showed that after 6-weeks the intervention group had significantly lower depression scores (MD -0.95, 95%CI -1.64, -0.27) and lower mindful awareness scores (MD -2.20, 95%CI -3.92, -0.48). Anxiety, mindful acceptance and medication adherence did not significantly change (anxiety MD 3.25, 95%CI -3.58, 1.07; mindful acceptance MD 0.79 95%CI - 0.66, 2.23; adherence MD -0.47, 95%CI -1.56, 1.47). After 6-weeks the control group had higher depression scores (MD 0.73, 95% CI 0.07, 1.39) and anxiety scores (MD 1.34, 95% CI 0.48, 2.20) but there was no significant change in any other measure change (mindful awareness MD -0.42 95%CI 2.09, -1.26; mindful acceptance MD 0.17, 95%CI -0.87, 1.21; adherence MD -0.07, 95%CI -1.83, 1.68).

Similarly, within-groups analysis after 3 months the intervention group had significantly lower depression than at baseline (MD -1.46, 95% CI -2.12, -0.81), significantly lower mindful awareness scores (MD -4.65, 95% CI -6.19, -3.10) and trend to lower anxiety scores that was not significant (MD -0.66, 95% CI -1.36, 0.04). There was no significant change in mindful acceptance (MD 0.66 95%CI -0.75, 2.07) or medication adherence (MD 0.15 95%CI -1.32, 1.63). The control group had significantly lower mindful awareness scores (MD -2.49, 95% CI -4.44, - 0.54) but no change in other measures (anxiety MD 0.47 95%CI -0.47, 1.40; depression MD 0.54 95%CI -0.14, 1.20; mindful acceptance MD 0.07 95%CI -1.55, 1.69; adherence MD -1.14 95%CI - 2.96, 0.68).

### Exploratory Analysis

#### Comparing ‘engaged’ participants in the intervention group vs. control

Exploratory analysis compared asthma quality of life in ‘engaged’ participants in the intervention group ie. those who accessed the intervention at least once (N = 65) vs. control participants. Between groups comparisons (correcting for baseline differences) showed higher 6-month AQLQ scores in the engaged group (MD 0.27, 95%CI -0.01, 0.54) but this was not significant. ACQ scores at 6-week follow-up were significantly higher in the intervention group (MD 0.37, 95%CI 0.14, 0.59). Similarly, at 3-months the intervention group had non-significantly better quality of life (MD 0.20, 95%CI -0.09, 0.49) and asthma control (MD -0.24, 95%CI -0.52, 0.04).

#### Group differences in minimal clinically important change (MCID)

Individual subject changes in AQLQ scores from baseline were assessed according to the achievement of MCID (0.5). The proportion of participants who had greater and less than MCID change in AQLQ is presented in Figure 2. A greater proportion of participants in the intervention group (6 weeks: 35%, 3 months: 43%) showed a relevant improvement in quality of life than in the control group (6 weeks: 22%, 3 months: 29%), and a higher percentage of control group (6 weeks: 17%, 3 months: 22%) than intervention (6 weeks: 11%, 3 months: 10%) showed a relevant decrease in quality of life.

**Figure 2:**
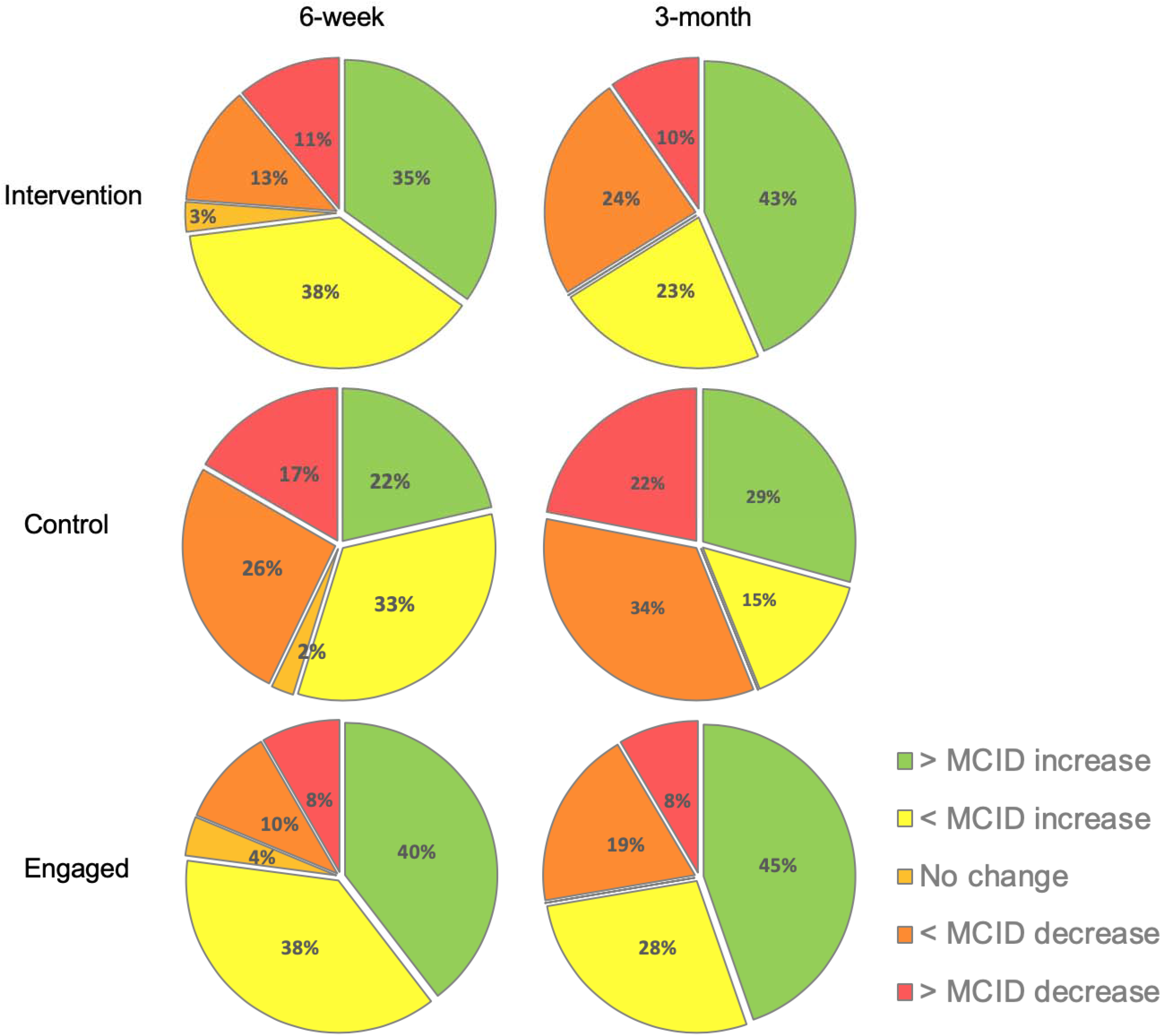
Change in primary endpoint (AQLQ) relative to MCID.

Using the reccommeded analysis described by Guyatt et al. (Guyatt et al., 1998), the number needed to treat (NNT) for one subject randomised to active arm to achieve a relevant improvement in quality of life above control was 4.88 (see Table 4).

**Table 4.** Number need to treat at 3-month followup (all patients who completed baseline and 3-month data).

|  |  | Intervention Group |  |  |
| --- | --- | --- | --- | --- |
|  |  | Improved<br>>MCID | Unchanged | Deteriorated >MCID |
| **Control**<br>**Group** | Improved<br>>MCID | 0.11 | 0.11 | 0.02 |
|  | Unchanged | 0.28 | 0.3 | 0.06 |
|  | Deteriorated<br>>MCID | 0.06 | 0.07 | 0.01 |
| **Proportion who received a benefit:** |  |  |  | 0.2 |
| **NNT:** |  |  |  | 4.88 |

Logistic regression analysis was used to compare the proportion of participants achieving an improvement greater than MCID after controlling for baseline AQLQ score. At 6-weeks, the those in the intervention group were non-significantly more likely to achieve a relevant improvement (OR 1.86, 95% CI 0.71, 4.86), with similar results at 3-months (OR 1.36, 95% CI 0.52, 3.58).

## Discussion

This pragmatic, randomised feasibility trial shows that the digital mindfulness intervention ‘Headspace’ is relevant and acceptable to at least a proportion of people with asthma, with the potential to benefit patients, so merits a fully-powered confirmatory RCT. Recruitment targets were achieved, and retention rates were comparable with previous randomized controlled trials of digital interventions in similar populations (McLean et al., 2016). Our sample ranged from 18 to 90 years old, demonstrating the potential utility of digital interventions to reach a broad patient demographic. Our questionnaire response rates were in line with previous relevant research (Morrison et al., 2016, Ainsworth et al., 2019) although it should be noted that using both internet and postal measures meant that some patients did not complete all measures (providing valuable information for a future trial).

Our primary outcomes of interest – asthma quality of life and asthma control – both demonstrated substantial improvement in the intervention group above baseline values, and consistent trends to improvements over the control group. A greater proportion of participants in the intervention group demonstrated an improvement of the minimally important clinical difference in asthma-specific quality of life, with a low number needed to treat of below 5 for a patient to experience a relevant improvement in asthma control. We also observed a trend to more positive anxiety and depression scores at follow-up in comparison to the control group. Although this feasibility study was not powered to evaluate differences between groups, our study consistently found promising trends to better outcomes for those in the intervention group compared to the control group. Importantly, we did not find evidence of change in medication adherence to explain the improvements obseved, consistent with the notion that MBTs may act as an adjunct intervention to standard pharmacological treatments to improve quality of life.

One of the strengths of the study was that engagement with Headspace exceeded other similar digital interventions in primary care patients with asthma (McLean et al., 2016). This suggests that this digital mindfulness intervention may be acceptable and accessible for many people with asthma. However, some participants did not use the intervention at all, suggesting that intervention reach could still be improved, and that this intervention may not be acceptable to all. Of those who did engage, the range of usage was large: some used it once or twice during the entire study whilst others used it several times a day. This is in line with cutting edge theories of digital behaviour change interventions: accessible interventions should be flexibly designed to allow for different usage patterns to suit individuals who may engage with behaviours in a variety of different ways that suit them (Ainsworth et al., 2017). In this study, we did not advise how frequently participants should use Headspace, nor if they should access specific components. However, detailed usage data was regarding specific Headspace practices that were preferred by individuals (eg ‘Managing Anxiety’ and ‘Stress’) which will inform the development of ‘Asthma-Specific’ Headspace guidance in a future trial. We also note that the exploratory nature of the feasibility study meant that we did not integrate our data-gathering platform (Lifeguide) with the intervention platform (Headspace) and therefore required participants to sign up to each individually streamlining these for a full trial would likely result in even more effective engagement. We also suggest using theory- and person-based approach to further maximise acceptability and effectiveness (Yardley et al., 2015).

As well as the encouraging results, this study had a sufficient sample size to support confidence in the exploratory findings. While the 2:1 randomisation process allows increased variability in the control group, it generated detailed intervention usage data that will inform a further full trial. The online nature of the study meant that study recruitment was particularly cost-effective and facilitated rigid study procedures, with very little possibility for researcher bias or protocol deviation).

There are several limitations to this study which must be acknowledged. Firstly, baseline comparisons indicated that those randomised to the intervention group tended towards impaired quality of life compared to the control group at baseline. Although our primary analysis controlled for these different baseline values, participants in the control group may have experienced a ceiling effect, and consequently had a reduced magnitude of improvement.

Although our findings suggested benefits of the intervention at 6-weeks and 3-months, this study does not explore long-term evidence of benefit (eg. for over a year). While it is possible that the benefits of mindfulness practice accrue over time, it may be that initial levels of engagement with the digital intervention ‘drop off’ as good habits formed by participants subside. Indeed, the inconsistent and heterogenous nature of asthma symptoms mean that other digital interventions have included specific content to remind users to re-engage when symptoms appear ((Ainsworth et al., 2019a)) and this should be included in a larger randomised controlled trial.

A complex behavioural intervention such as mindfulness means that participants are not blind to their group allocation. However, psychological benefits to receiving a treatment are, in the case of mindfulness, fundamental treatment components that should be included in evaluation (Ainsworth et al., 2019b) and therefore we consider our pragmatic feasibility trial an effective design, especially given the remote nature of the study (ie. Researcher blinding could not be an issue).

Of more concern is the consideration of ‘reach’: that patients with the most impaired quality of life and asthma control (who are likely to benefit most from adjunct therapies) may not be willing to sign up to digital interventions, particularly treatments such as mindfulness. While mindfulness is increasingly common in the public sphere (and therefore increasingly acceptable; (Kachan et al., 2017)), and internet access is more widespread, care must be taken not to entrench digital inequality (Hargittai et al., 2018). Therefore, any further research must use theory- and person-based approaches to ensure that a full trial and subsequent dissemination is accessible across as broad a socio-economic demographic as possible.

The remote nature of the study also meant that we were unable to measure objective, physiological markers of asthma, such as lung function and health-resource use. Although evidence suggests that subjective self-report is a more accurate predictor of quality of life (Janssens et al., 2012), such mechanisms should not be overlooked in order to understand the mechanisms by which the observed improvements in asthma control and asthma quality of life occurred. Understanding the mechanisms of psychological and behavioural treatments is important to determine whether the benefits of such treatments are ‘non-specific’ (see (Ainsworth et al., 2019b) or target specific psychological mechanisms that may be dysfunctional in patient groups. Mindfulness, which advocates a non-judgemental awareness of thoughts and feelings, may lead to better asthma outcomes through improved symptom perception – which has previously been demonstrated to be a better predictor of quality of life for people with asthma than objective lung function impairment (Janssens et al., 2012). Similarly, mindfulness may improve illness perceptions (beliefs and emotional responses to their condition) – which subsequently effect a range of asthma outcomes including disease management (Kaptein et al., 2010). That we did not observe changes in medication adherence suggests that mindfulness could be an effective adjunct therapy to existing pharmacological treatment (especially when delivered digitally, in a flexible format that can be tailored for the heterogenous population of adults with asthma).

## Conclusion

This study demonstrated the feasibility of a digital mindfulness intervention for people with asthma in primary care, suggesting benefits for asthma control and quality of life, anxiety and depression. The intervention was acceptable to patients, although engagement levels varied across the sample. With appropriate modification of trial procedures, these data support the feasibility of a confirmatory randomised-controlled trial.

## Data Availability

The analysis scripts used during the current study are available in the FIGSHARE repository. The data is available in line with the consent collected from participants; by request.

## Declarations

### Declaration of Interests

The authors declare the following financial interests/personal relationships which may be considered as potential competing interests:

BA has received fees for speaking at an educational meeting hosted by Astra Zeneca, and an expert advisory panel Roche.

MT has received speaker’s honoraria for speaking at sponsored meetings or satellite symposia at conferences from the following companies marketing respiratory and allergy products: GSK, Novartis. He has received honoraria for attending advisory panels with; Boehringer Inglehiem, GSK, Novartis,. He is a recent a member of the BTS SIGN Asthma guideline steering group and the NICE Asthma Diagnosis and Monitoring guideline development group.

RD report personal fees from Synairgen, which is outside the submitted work. RD also reports receiving fees for lectures at symposia organized by Novartis, AstraZeneca and TEVA, consultation for TEVA and Novartis as member of advisory boards, and participation in a scientific discussion about asthma organized by GlaxoSmithKline.

BS, DR, ML and SS have no conflicts of interest.

### Ethics

Ethical approval was given by the South Central Hampshire Research Ethics Committee (17/SC/0088) and the University of Southampton Ethics and Research Governance Online (24515). The study was in line with The Code of Ethics of the World Medical Association (Declaration of Helsinki). All participants provided informed consent before taking part.

### Funding

The study was supported by a research project grant from the NIHR School of Primary Care (FR13:373). During this project BA was supported by an NIHR Post-doctoral Translational Fellowship from the NIHR School of Primary Care. The funding body had no input into design, analysis or interpretation of data, nor any input into the manuscript. The views expressed are those of the authors and not necessarily those of the NIHR, MRC, the NHS or the Department of Health.

Headspace Inc. provided free access to their intervention for participants in the study. They had no input into design, analysis or interpretation of data, nor the decision to submit for publication, although were asked to provide information for the manuscript.

### CRediT Author Statement

**BA:** Conceptualization, Methodology, Software, Formal analysis, Investigation Writing - Original Draft, Writing - Review & Editing, Visualisation, Funding acquisition. **SS:** Methodology, Investigation, Writing - Review & Editing. **BL**: Methodology, Formal analysis, Writing - Review & Editing, Funding acquisition. **DR:** Methodology, Resources, Writing - Review & Editing. **ML:** Data Curation, Investigation, Writing - Review & Editing. **RD:** Conceptualization, Writing - Review & Editing, Supervision. **MT:** Conceptualization, Methodology, Writing - Review & Editing, Supervision, Funding acquisition.

## Abbreviations

ACQ: Asthma Control Questionnaire
AQLQ: Asthma Quality of Life Questionnaire
GP: General Practitioner
HADS: Hospital Anxiety and Depression Scale
MARS-A: Medical Adherence Report Scale
MBIs: Mindfulness-based Interventions
MBCT: Mindfulness-based Cognitive Therapy
MBSR: Mindfulness-based Stress Reduction
MCID: Minimum clinically important difference
MD: Mean Difference
NIHR: National Institute of Health Research
NNT: Number needed to treat
OR: Odds ratio
PHLMS: Philadelphia Mindfulness Scale
RCT: Randomised Controlled Trial

## Notes

### Clinical Trial

ISRCTN52212323

